# The effects of vaccination on the disease severity and factors for viral clearance and hospitalization in Omicron-infected patients :A retrospective observational cohort study from recent regional outbreaks in China

**DOI:** 10.1101/2022.06.28.22276985

**Authors:** Hongru Li, Xiongpeng Zhu, Rongguo Yu, Xin Qian, Yu Huang, Xiaoping Chen, Haibin Lin, Huiming Zheng, Yi Zhang, Jiarong Lin, Yanqin Deng, Wen Zhong, Yuejiao Ji, Qing Li, Jiabin Fang, Xiaojie Yang, Rong Lin, Sufang Chen, Zhijun Su, Baosong Xie, Hong Li

## Abstract

**Object:** It remains unelucidated regarding the effects of vaccination on disease severity and factors for viral clearance and hospitalization in omicron-infected patients.

**Methods:** The clinical manifestations of 3,265 Omicron-infected patients (BA.2 variant; the Omicron group) were compared with those of 226 Delta-infected patients (the Delta group).A Multi-class logistic regression model was employed to analyze the impacts of vaccination doses and intervals on disease severity; a logistic regression model to evaluate the risk factors for hospitalization; R 4.1.2 data analysis to investigate the factors for time for nucleic acid negativization (NAN).

**Results:** Compared with the Delta group, the Omicron group reported a fast transmission, mild symptoms, and lower severity incidence, and a significant inverse correlation of vaccination dose with clinical severity (OR: 0.803, 95%CI: 0.742-0.868, p<0.001). Of the 7 or 5 categories of vaccination status, the risk of severity significantly decreased only at ≥21 days after three doses (OR: 0.618, 95% CI: 0.475-0.803, p<0.001; OR: 0.627, 95% CI: 0.482-0.815, p<0.001, respectively). The Omicron group also reported underlying illness as an independent factor for hospitalization, sore throat as a protective factor, and much shorter time for NAN [15 (12,19) vs. 16 (12,22), p<0.05]. NAN was associated positively with age, female gender, fever, cough, and disease severity, but negatively with vaccination doses.

**Conclusion:** Booster vaccination should be advocated for COVID-19 pandemic-related control and prevention policies and adequate precautions should be taken for patients with underlying conditions.

## INTRODUCTION

The COVID-19 outbreak in early 2020 has brought about a global pandemic, with a toll of tens of thousands of deaths (6,294,252 in total as of May 30, 2022). Of the new variants of the novel coronavirus, the Omicron mutant was first identified in Botswana on November 9, 2021 and soon spread worldwide from South Africa.^1^ It was imported to Hong Kong and Chinese mainland Tianjin in November 2021, and has spread to many regions. The BA.2 variant has infected over 600,000 people in Shanghai^2^ and a total of 3,265 patients in Quanzhou.

Compared with the Delta variant, the Omicron variant features a higher transmissibility (R0 value: 9.5 vs. 3.4)^3^ and a shorter incubation period (3.6 d vs. 4.6 d)^4^. It is also characterized by more noticeable upper respiratory symptoms (strong nasal tropism) and lower incidence of pneumonia (less obvious lung tropism).^5^ Due to the decreased virulence, the Omicron variant infection displays milder clinical manifestations, reduced hospitalization risk (about 50-70% of that of the Delta variant),^6^ and decreased mortality rate (about half of the Delta variant)^7^.

To date, vaccination provides effective protection^8^ against the Omicron variant and reduces hospitalization risk. However, mutations in S protein compromise the effectiveness of neutralizing antibodies and vaccination, leading to immune escape and breakthrough infections.^9,10^ Given that booster vaccines can only reinforce the protection temporarily, ^8^ the COVID-19 pandemic is far from over. A South African study has predicted 100 million infections on the 100th day of Omicron prevalence, 160,000 deaths and a case fatality rate of about 0.58%.^7^ WHO has recently announced that the Omicron strain has caused 830,000 excess deaths.^11^ The outbreak of Hong Kong Omicron epidemic from January to March 2022 has claimed 9,115 deaths, with a mortality rate of 0.76%.^12^ Therefore, the gravity of the Omicron variant should not be underestimated and it remains urgent to formulate effective epidemic prevention strategies.

Currently, little relevant literature is available to elucidate the effects of vaccination on the disease severity of the Omicron variant infection and the hazard factors for hospitalization and time for viral clearance.^13^ Therefore, the current study tackled this very issue by comparing two recent regional COVID-19 surges in China (one local Omicron and one local Delta outbreak), so as to provide evidence for drafting effective public health policies in coping with the pandemic.

## METHOD AND OBJECTS

### Study design and study subjects

The flow chart of the study design can be found in the supplementory data(**FigureS1**).

All the SARS-Cov-2 cases with positive nucleic acid during two recent regional outbreaks in Fujian, China (from 2021 to the present) were confirmed by fluorescence real-time PCR (RT-PCR) during two recent regional outbreaks in Fujian, China from 2021 to the present. They were divided into the Omicron group: 3,265 Omicron BA.2-infected patients in Quanzhou City, Fujian, China between March 13 and May 6, 2022; and the Delta group^14^: 226 Delta-infected patients (Delta B.1.167.2) in Putian City, Fujian, China from September 10 to October 20, 2021. Different from previous foreign studies^15^ that only included symptomatic populations, this study enrolled all nucleic acid-positive cases in the regional outbreaks, including asymptomatic and mild patients. Informed consent was granted by all patients and the study protocol was approved by the Ethics Committee of the First Hospital of Quanzhou City (No. 202212) and of the Affiliated Hospital of Putian University (No. 202152).

### Data involvement

Clinical and experimental data were collected from electronic medical records using standardized data collection forms. The clinical data of the patients included symptoms, signs, and treatment prognosis. The laboratory data included the cycle threshold (CT) value of SARS-Cov-2 RT-PCR upon admission and serological results of whole blood cells, biochemical tests, hemagglutination function, C-reactive protein, and IL-6 within 24 hours after admission(Test kits and apparatus were described in **TableS1**,normal value of test results were shown in **TableS2)**. The information of vaccination time and doses of the patients was obtained from Chinese Health Organization, most patients(99.47%) received inactivated vaccination (including Sinopharm and CoronaVac vaccine), and a few(0.46%) received Recombinant novel coronavirus protein vaccine (Zhifei Longcom) and 0.07% received mRNA vaccine (CanSino). Nasopharyngeal swabs were collected every day from the 7th day of the disease course until the nucleic acid negativization (NAN). The time for NAN was defined as the beginning of the patient’s disease course or the day of a positive SARS-Cov-2 neucleic acid swab test to the time of two consecutive negative nucleic acid swab tests that were obtained at an interval of more than 24 hours. The NAN standard was defined as a CT value of over 35 in the Omicron group^16^ and of over 40 in the Delta group^14^.

### Clinical management

The disease severity was classified into asymptomatic, mild (no pneumonia on chest X-ray), moderate (pneumonia by chest imaging), severe (SpO_2_<93%, supplemental oxygen needed) or critical (requiring RICU or mechanical Ventilation) categories. In the Omicron group, the asymptomatic and mild patients were quarantined in the mobile cabin hospital (MCH) while patients with moderate or severer symptoms were hospitalized for treatment (as the latter group of patients were hospitalized and there were few severe/critical cases, patients with moderate symptoms were recruited for the analysis of hospitalization risks). All Delta patients were treated in hospitals. Most patients in the Omicron group received traditional Chinese medicine (TCM) treatment and were monitored for vital signs, body temperature and respiration. Oxygen inhalation treatment was prescribed for patients with SpO_2_ below 93%; nematevir/ritonavir antiviral treatment was administered to patients showing unstable disease progression within 3-5 days and no contraindications; specific immunotherapies, such as Covid-19 immunoglobulin, neutralizing antibody or convalescent plasma, were initiated for high risk patients with high viral load or a rapid disease progression in the early stage of the disease course; glucocorticoids were only used for people with specific needs and antibiotics were for patients complicated with bacterial infection.^16^ The Delta group received the same treatment schemes, except for Nematevir/Ritonavir, which was not yet available then.^14^

### Statistics analysis

For descriptive analysis, we presented data as mean (variance) or median [interquartile range (IQR)] for continuous parameters and as frequency (percentage) for categorical variables using IBM SPSS Stastistic25 data analysis software. Wilcoxon rank sum testing was employed for numerical variables and Fisher’s precision probability test was adopted for categorical variables, if a sample size was less than 5 in the cells of the contingency table of the corresponding indicators, and the Chi Square test was used for the rest. An ordered multivariate logistic regression model was used to analyze the effects of vaccine doses and intervals on disease severity of patients.A disordered multi-category logistic regression model to analyze the hospitalization risk factors.The factors affecting the time for NAN which illuminated the viral clearance was examed by the least-squares regression of the R 4.1.2 data analysis. A two-tailed P value of less than 0.05 was considered statistically significant.

## RESULT

### Baseline information

The demographic characteristics of the patients are shown in **Table 1**. The total number of infected persons in the Omicron group and the Delta group was 3,265 and 266, respectively. The median age of the Omicron-infected patients was higher than that of the Delta-infected patients [36 (25,48) vs. 32 (9,46)]^14^; the most common primary diseases in both groups were hypertension (5.61% vs. 2.65%) and diabetes (2.62% vs. 4.87%); 16 pregnant women were found in the Omicron group.

**Table 1.**
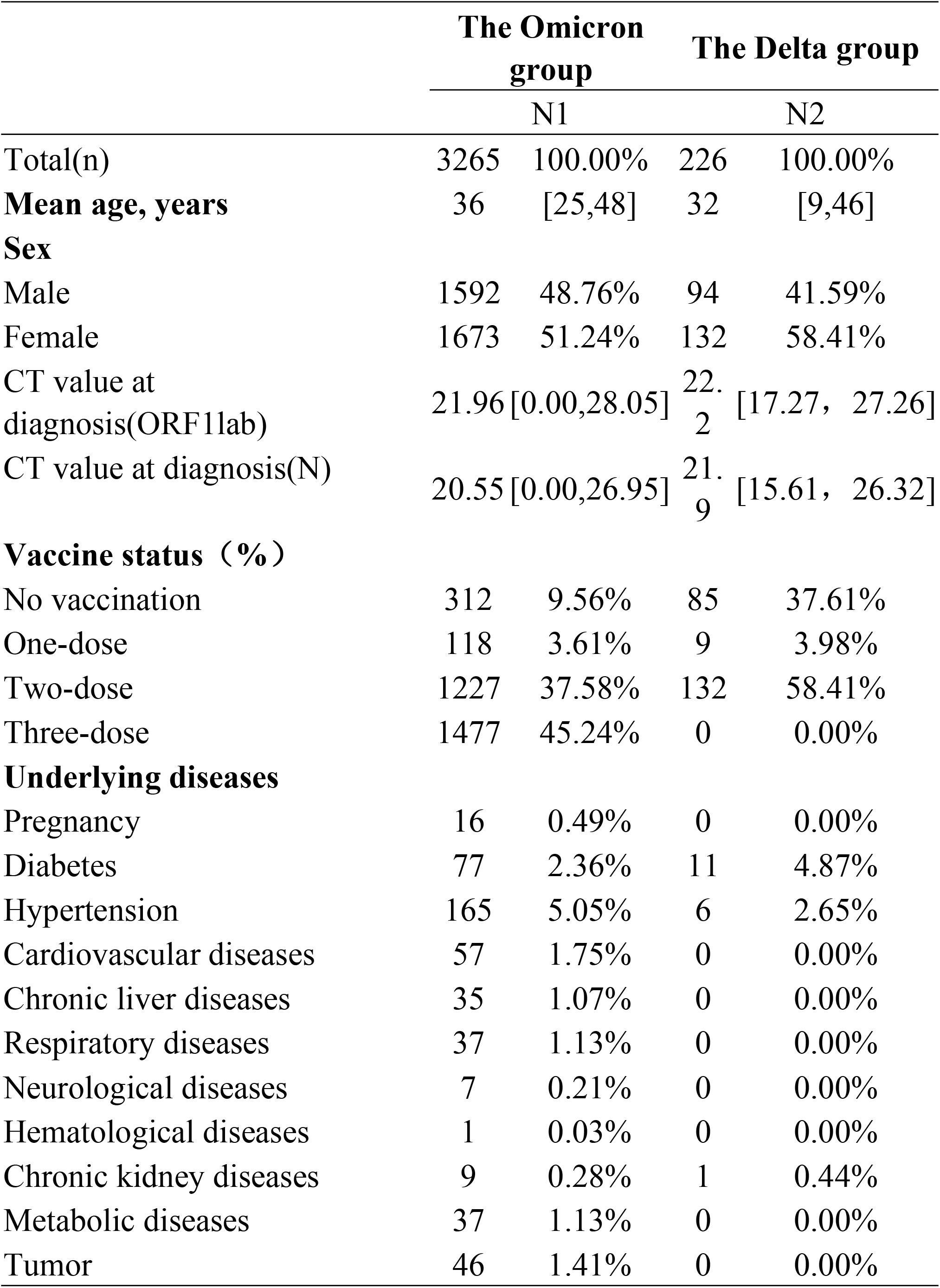
Baseline information of the Omicron and Delta group.

Due to the specific vaccination strategy and policies in China, in the Omicron group, children aged 0-2 years old were not vaccinated while patients under 12 years old were mostly vaccinated with two doses (53.09%) and those over 12 years old were mostly vaccinated with three doses (49.23%). In the Delta group, none of the children under 12 years old was vaccinated while most of the patients older than 12 years were vaccinated with two doses (88.59%) **(Table 1)**.

### Comparison of clinical characteristics between Omicron-infected patients and Delta-infected patients

Compared with the Delta group, the Omicron group reported fewer severe symptoms (p<0.05), including asymptomatic (61.13% vs. 2.21%), mild (31.82% vs. 36.73%), moderate (2.76% vs. 58.41%), severe (0.06% vs. 1.77%) and critical (0.03% vs. 0.88%) categories, respectively, with the incidence of severe critical symptoms paralleling the increasing age **(Table 2)**. Furthermore, the Omicron group showed significantly lower rates of pneumonia (2.85% vs. 59.73%, p<0.001) and respiratory failure (0.06% vs. 2.65%, p<0.001), and less ICU occupancy (0.09% vs. 2.21%, p<0.05) **(Table 2)**. No deaths were reported in both groups. Due to the imbalance of disease severity in the Omicron infection, we used R 4.1.2 to randomly to select 90 asymptomatic and mild samples respetively from the asymptomatic and mild patients in the Omicron group respectively, forming a new data set of 270 cases together with another 90 moderate cases.The comparison of the data of the 270 Omicron cases and 220 Delta cases revealed that compared with the Delta patients, the Omicron patients were more prone to pharyngeal pain (OR: 15·62; 95%CI: 5·29-61·9),and headache or muscle pain (OR: 6·23; 95% CI: 2·63-15·84), and less prone to fatigue (OR: 0·28; 95% CI: 0·15-0·50, p < 0·0001) and fever (OR: 0·14; 95% CI: 0·09-0·22, p < 0·001)**(Figure S2)**

**Table 2.** Comparison of clinical characteristics between the Omicron and Delta group

|  | Omicron group |  | Delta group |  | p-value |
| --- | --- | --- | --- | --- | --- |
| Total(n) | 3265 |  | 226 |  |  |
| Vaccination | 2822 | 86.43% | 137 | 60.62% | <0.001 |
| Number of vaccine doses |  |  |  |  | <0.001 |
| No vaccination | 312 | 9.56% | 85 | 37.61% |  |
| One dose | 118 | 3.61% | 9 | 3.98% |  |
| Two doses | 1227 | 37.58% | 132 | 58.41% |  |
| Three doses | 1477 | 45.24% | 0 | 0.00% |  |
| Clinical features |  |  |  |  |  |
| CT value at diagnosis(ORF1lab) | 21.96 | [0.00,28.05] | 22.20 | [17.27, 27.26] | 0.124 |
| CT value at diagnosis(N) | 20.55 | [0.00,26.95] | 21.90 | [15.61, 26.32] | 0.063 |
| Fever n (%) | 436 | 13.35% | 147 | 65.04% | <0.001 |
| Nasal congestion n (%) | 30 | 0.92% | 3 | 1.33% | 0.470 |
| Sore throat n (%) | 395 | 12.10% | 4 | 1.77% | <0.001 |
| Cough n (%) | 770 | 23.58% | 80 | 35.40% | <0.001 |
| Expectoration n (%) | 259 | 7.93% | 42 | 18.58% | <0.001 |
| Dyspnea n (%) | 26 | 0.80% | 3 | 1.33% | 0.431 |
| Headache or Sore muscle n (%) | 220 | 6.74% | 13 | 5.75% | 0.679 |
| Fatigue n (%) | 323 | 9.89% | 64 | 28.32% | <0.001 |
| Inappetence n (%) | 94 | 2.88% | 8 | 3.54% | 0.538 |
| Diarrhea n (%) | 33 | 1.01% | 7 | 3.10% | 0.017 |
| Disease severity |  |  |  |  | <0.001 |
| Asymptomatic | 1996 | 61.13% | 5 | 2.21% |  |
| Mild | 1039 | 31.82% | 83 | 36.73% |  |
| Moderate | 90 | 2.76% | 132 | 58.41% |  |
| Severe | 2 | 0.06% | 4 | 1.77% |  |
| Critical | 1 | 0.03% | 2 | 0.88% |  |
| Laboratory diagnostics |  |  |  |  |  |
| WBC count (10^9/L)average(SD) | 5.1 | [3.86,6.84] | 6.25 | 2.01 | <0.001 |
| Lymphocyte count (10^9/L)Median(IQR) | 1.56 | [1.09,2.26] | 1.46 | [1.02,2.22] | 0.132 |
| APTT (s) | 32.3 | [30.00,35.00] | 28.00 | [23.98,31.35] | <0.001 |
| D-D (mg/L) | 0.27 | [0.17,0.47] | 0.31 | [0.18,0.53] | 0.050 |
| CRP<5mg/L | 815 | 24.96% | 157 | 69.47% | <0.001 |
| IL-6(pg/ml) | 6.74 | [3.12,32.50] | 7.00 | [3.82,13.98] | 0.466 |
| LDH (U/L) | 199 | [164,236] | 223.10 | 65.53 | <0.001 |
| Treatment and prognostic indicator |  |  |  |  |  |
| Pneumonia n(%) | 93 | 2.85% | 135 | 59.73% | <0.001 |
| respiratory failures,n(%) | 2 | 0.06% | 6 | 2.65% | <0.001 |
| Nucleic acid negativization time(days),Median(IQR) | 15 | [12,19] | 16.00 | [12.00,22.00] | <0.001 |
| ICU occupancy | 3 | 0.09% | 5 | 2.21% | <0.001 |
| Intubation rate,n(%) | 1 | 0.03% | 2 | 0.88% | 0.004 |
| TCM treatment,n(%) | 2973 | 91.06% | 214 | 94.69% | 0.046 |
| Antibiotic usage rate,n(%) | 19 | 0.58% | 15 | 6.64% | <0.001 |
| Immunotherapy,n(%) | 4 | 0.12% | 42 | 18.58% | <0.001 |
| Antiviral drug | 31 | 0.95% | 0 | 0.00% | —— |
| Hormones therapy,n(%) | 0 | 0.00% | 2 | 0.88% | —— |
| Death rate,n(%) | 0 | 0.00% | 0 | 0.00% | —— |

Note: For descriptive analysis, data were expressed as mean (variance) or median [interquartile range (IQR)] for continuous parameters and as frequency (percentage) for categorical variables. For numerical variables, the Wilcoxon rank sum testing was used; for categorical variables, Fisher's precision probability test was employed if a sample size was less than 5 in the cells of the contingency table of the corresponding indicators, and the Chi Square test was adopted for the rest by IBM SPSS Stastistic25 data analysis software.The total sample size of Omicron infected group was 3265 cases, and the number of serological examination sample was 1195 cases.

### Impacts of vaccination on disease severity of the Omicron infection

The results showed that disease severity was markedly reduced by the increased vaccination doses (OR: 0.803; 95%CI: 0.742-0.868, p<0.01)**(Table 3)**. The association between vaccination doses and intervals and disease severity was referred to the unvaccinated Omicron-infected individuals.. The vaccination status was divided into seven categories (No vaccination,<21 days post dose one,≥21 days post dose one, but no dose two,<21 days post dose two,≥21 days post dose two, but no dose three,<21 days post dose three,≥21 days post dose three, **(Figure1A, Table S3)**) or five categories (No vaccination,<21 days post dose one,≥21 days post dose one to <21 days post dose two,≥21 days post dose two to <21 days post dose three,≥21 days post dose three,**(Figure1B,TableS4)**).). The analyses showed that regardless of the 7 or 5 categories of vaccination status, the disease severity was significantly reduced only at ≥21 days after 3 doses (OR: 0.618; 95% CI: 0.475-0.803, p<0.001; OR: 0.627, 95% CI: 0.482-0.815, p<0.001 respectively), with no significant difference found for the other categories **(Figure1A-1B)**. ^17^

**Figure 1:**
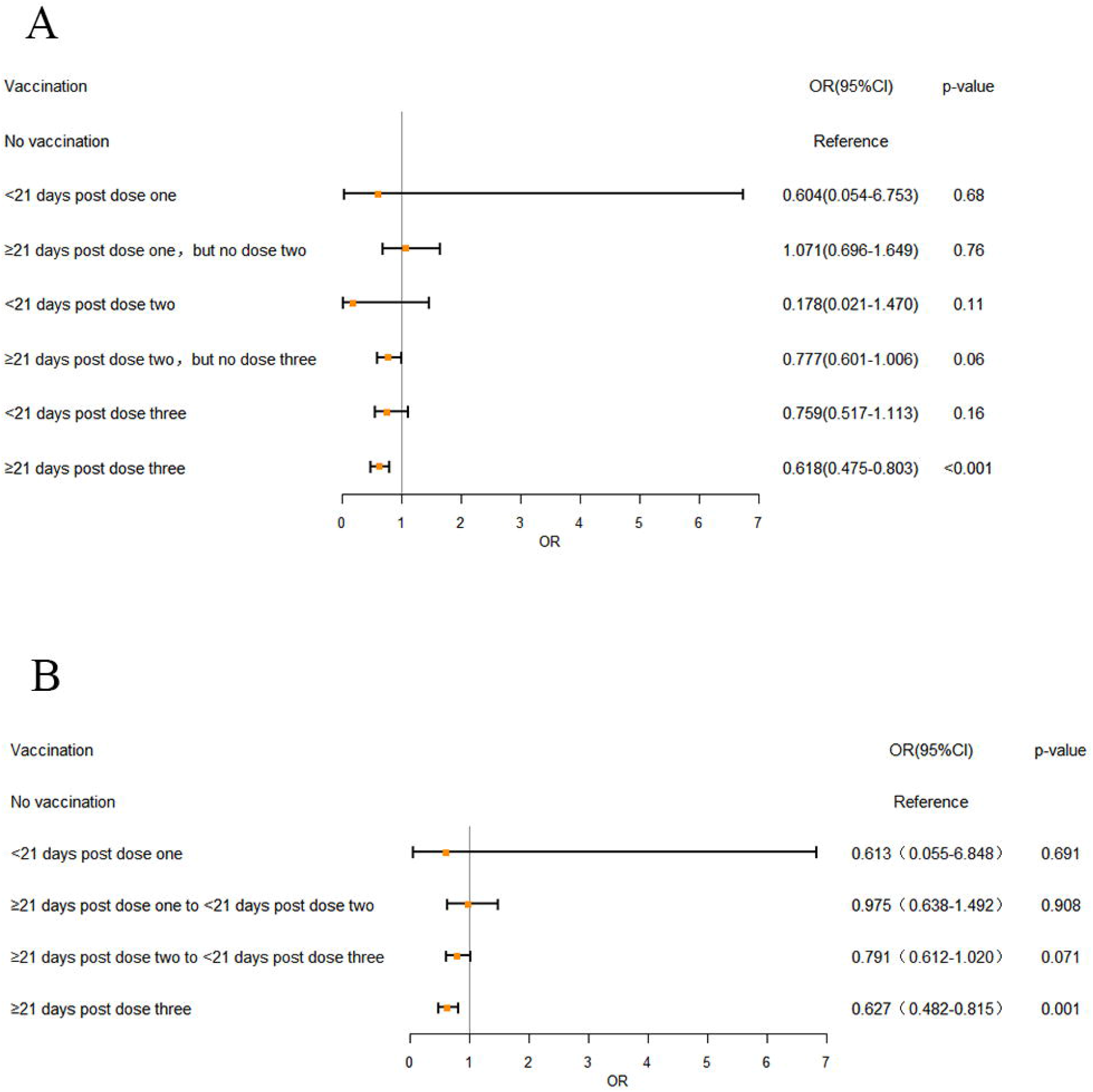
The effects of different vaccination doses and intervals on disease severity in Omicron infection. (A) 7 different vaccination doses and intervals, (B) 5 different vaccination doses and intervals. OR: odds ratio. Error bars represent 95% CI. Unvaccinated individuals infected with Omicron served as the reference category. A multivariate ordered logistic regression model was used to analyze the effects of vaccination doses and intervals on disease severity of the patients. Doses of vaccines were treated as independent variable and three clinical symptom categories, “asymptomatic”, “mild” and “moderate”, were treated as dependent variables and designated to indicate the increasing disease severity, with 3 severe and critical patients excluded from analyses. The analyses showed that regardless of the 7 (A) or 5 categories (B) of vaccination status, the disease severity was significantly reduced only at ≥21 days after 3 doses (OR: 0.618; 95% CI: 0.475-0.803,p<0.001; OR: 0.627, 95% CI: 0.482-0.815, p<0.001 respectively), with no significant difference found for the other categories.

**Table 3.** Relationship between vaccination and disease severity of the Omicron infection.

| Parameter estimated value |  |  |  |  |
| --- | --- | --- | --- | --- |
|  |  | Coefficients | OR ( 95%CI ) | p-value |
| Threshold | Asymptomatic | 0.154 | 1.166(0.967-1.408) | 0.108 |
|  | Mild | 3.078 | 21.715(16.593-28.417) | <0.001 |
| Location | Vaccination doses | -0.219 | 0.803(0.742-0.868) | <0.001 |

Note: Multivariate ordered logistic regression model was used to analyze the effects of vaccine doses and intervals on disease severity of the patients. Three clinical symptom categories, “asymptomatic”, “mild” and “moderate”, were treated as dependent variables of vaccine doses and intervals and designated to indicate the aggravating disease severity, with 3 severe and critical patients excluded from analyses. The Results showed that the increased number of vaccines significantly reduced the risk of severe diseases (OR=0.803, 95%CI: 0.742-0.868, $p<0.001$ ).

### The time for nucleic acid negativization (NAN) and influencing factors in the Omicron group

The results showed that the Omicron group reported a much shorter NAN time when compared with the Delta group [15 (12,19) vs. 16 (12,22), p<0.05]**(Table 2)**. For the influencing factors, the NAN time of the Omicron group was negatively correlated with fever and disease severity, which was similar to that of the Delta group.^14^ Moreover, the NAN time was negatively correlated with female gender, age, and cough, but positively with vaccination, which was different from the Delta group **(Table 4)**.^14^ In the Omicron group, the CT value of nucleic acid on admission did not affect the time of NAN. The relationship between COVID-19 antibody and the time for NAN was not analyzed because the titer of COVID-19 antibody was not detected in the omicron surge.

**Table 4.** Analysis of influencing factors for the time for nucleic acid negativization (NAN) in Omicron infection.

|  | Univariable analysis |  | Multivariable analysis |  |
| --- | --- | --- | --- | --- |
|  | Coefficients | p-value | Coefficients | p-value |
| ( Intercept) |  |  | 14.6187 | <0.001 *** |
| Sex | 0.8651 | 0.002 ** | 0.9727 | <0.001 *** |
| Age | 0.0142 | 0.049 * | 0.0195 | 0.018 * |
| Fever | 0.7811 | 0.013 * | 0.7531 | 0.023 * |
| Cough | 0.6032 | 0.036 * | 0.5716 | 0.046 * |
| Underlying diseases | 1.2256 | 0.0129 * | 0.9887 | 0.062 . |
| **Vaccination** |  |  |  |  |
| One dose | -0.7353 | 0.266 | -0.5971 | 0.362 |
| Two doses | -0.9409 | 0.023 * | -0.9219 | 0.025 * |
| Three doses | -1.0845 | 0.009 ** | -1.2253 | 0.005 ** |
| **Disease severity** |  |  |  |  |
| Mild | 0.7419 | 0.011 * | 0.4506 | 0.140 |
| Moderate | 1.3319 | 0.035 * | 1.0660 | 0.091 . |
| Severe | 7.6349 | 0.025 * | 7.4921 | 0.026 * |
| IL-6 | 0.0002 | 0.238 |  |  |
| CRP<5mg/L | 0.2842 | 0.349 |  |  |
| PCT | -0.0096 | 0.847 |  |  |
| WBC count | -0.0023 | 0.404 |  |  |
| LDH | 0.0003 | 0.847 |  |  |
| Lymphocyte count | 0.0102 | 0.001 ** | 0.0102 | 0.002 ** |
| DDi | 0.0024 | 0.85 |  |  |
| APTT | -0.0110 | 0.688 |  |  |
| Special treatment | 0.7032 | 0.040 * | 0.6018 | 0.084 . |
| ORF1lab | 0.0130 | 0.305 |  |  |
| N | 0.0093 | 0.475 |  |  |

Note: Data from 1,195 patients with blood test results accessible were selected for analysis. The factors affecting NAN time in the Omicron infection were analyzed by least-squares regression of the R 4.1.2 data analysis. Indices of statistical differences in the univariate analysis were included in the multivariate logistic regression analysis.
NAN: nucleic acid negativization,
\*indicates that the p-value of the variable is less than 0.05.
\*\*indicates that the p-value of the variable is less than 0.01.
\*\*\*indicates that the p-value is less than 0.001.

### Risk factors for hospitalization in the Omicron group and the Delta group

The results showed that the underlying disease was the most significant independent risk factor(OR: 1.299, 95% CI: 0.601-2.811, p<0.05) and sore throat was a protective factor(OR:0.367, 95% CI: 0.16-0.84, p<0.05), with mild patients as the reference category **(Table 5)**. The above results were significantly different from those of the Delta group **(Table S5)**, in the later group, which an advanced age and cough were risk factors for the moderate patients while the two-dose vaccination acted as a protective factor.

**Table 5.**
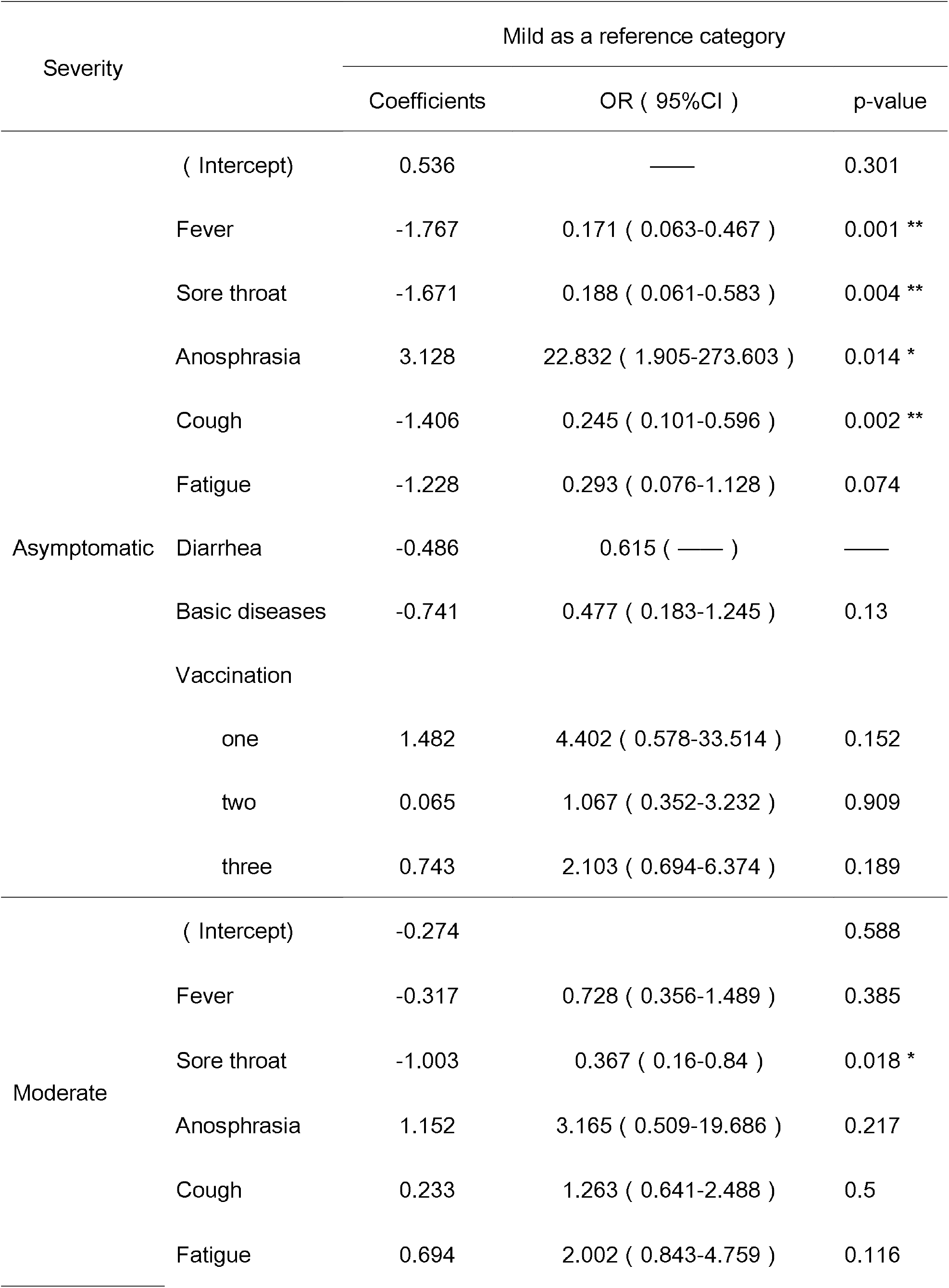
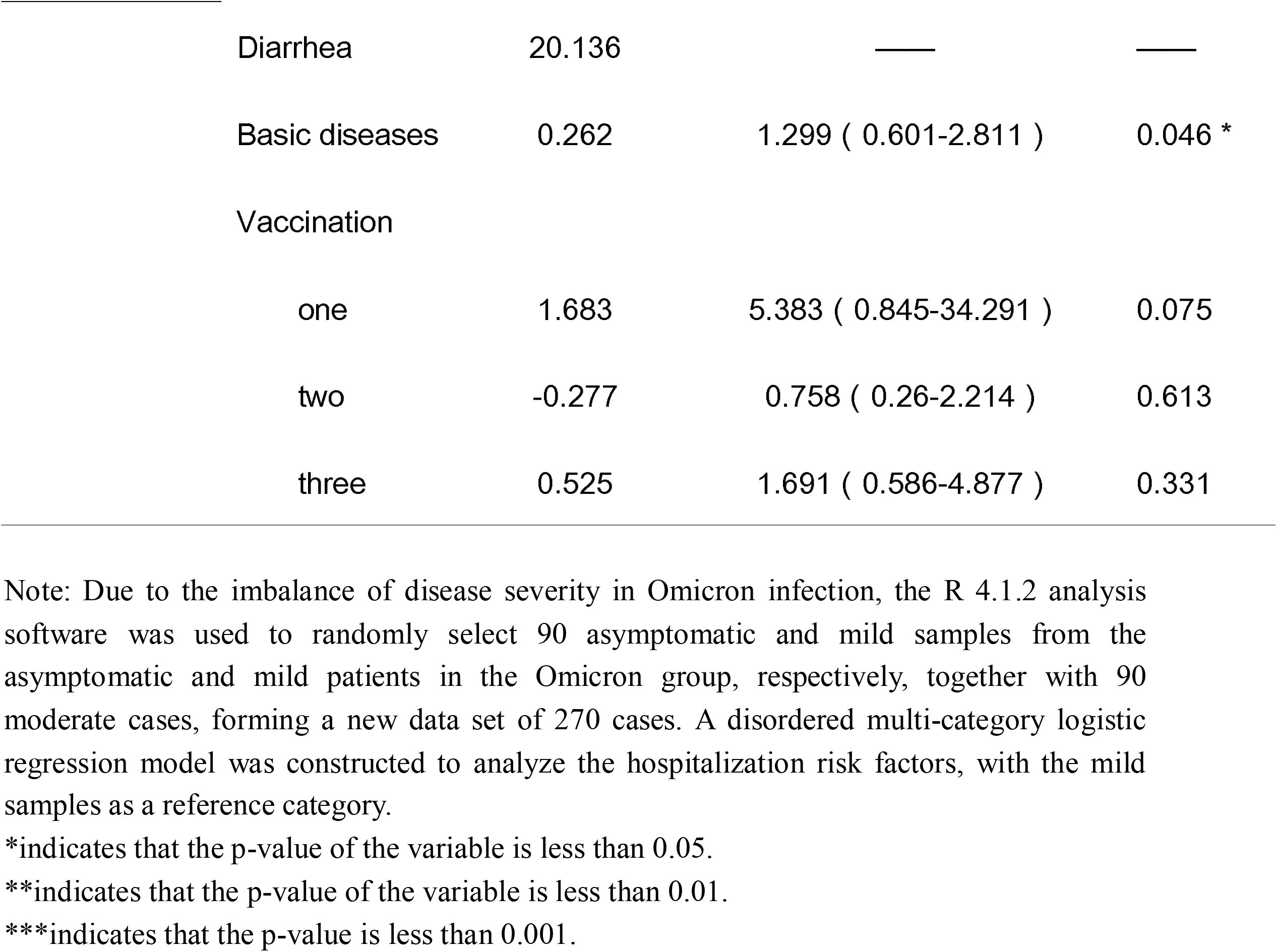
Analysis of hospitalization risk factors in Omicron infection.

## DISCUSSION

Omicron variants have become the dominant VOC in the world, including multiple subfamilies known as BA.1, Ba.2 and Ba.3, Ba.4, Ba.5.^1,18^ Compared with the Delta counterpart, the disease severity and mortality of the Omicron group decreased significantly. The data obtained in this study are consistent with the results of previous studies.^19^ The findings from the current study provide strong evidence for the timely adjustments to the pandemic control and prevention policies, especially for the developing countries that are challenged with a large population basis but inadequate healthcare resources.

Different countries have responded to the COVID-19 pandemic with vaccination, but it is still obscured whether vaccination is effective in reducing the transmission and severity of Omicron infection. Our study showed that despite the vaccination, many breakthrough infections still occurred, of which more than 45% of patients received a third booster vaccine and more than 86% of patients received two, which is consistent with previous studies.^9,10^ Of interest, the number of vaccinations, however, was significantly negatively correlated with the severity of Omicron infection, suggesting that vaccination can prevent the infection from exacerbation. Further analysis showed that compared with the two-dose regimen, which was found statistically ineffective in the current study, a booster vaccine fared much better and provided the strongest protection at ≥21 days. This finding is consistent with that of previous studies^8^ reporting the best effect at 2-4 weeks after the booster vaccination. A potential explanation may lie in that the booster vaccine restores the Omicron-weakened neutralization of the antibodies^8^ and enhances the protection by activating the specific T cell immunity^20^. In addition, the vaccination-induced memory B cells also play an important role in fighting against the Omicron variant.^21^ Therefore, the vaccination with a booster vaccine is of great significance in the prevention of the Omicron epidemic. As the effectiveness of the booster shots markedly wanes after 9 weeks,^8^ a fourth booster dose surfaces as a potential option in the response to the Omicron variant epidemic. Recent studies^22,23^ have produced inconsistent conclusions regarding the necessity of a fourth dose however. Meanwhile, the development of new vaccines^24^ against specific Omicron variants is currently under way in some countries, which may offer an important means to deal with the Omicron outbreak, but their effectiveness awaits further verification.

In the current study, the hospitalization risks in the Omicron group were defined as the factors in patients of the moderate clinical category who needed to be hospitalized. We found that underlying diseases were an independent risk factor and that sore throat was a significant protective factor. Pharyngeal pain, one of the manifestations of the upper respiratory tract infection, serves as one of the significant clinical manifestations of Omicron infection, which differs from those of Delta infection.^14^ The discrepancy may be related to the strong characteristic nasal tropism but less apparent lung tropism of the Omicron variant.^5^ In the current study, we found that, different from the Delta group,^14^ the underlying disease was an independent risk factor for hospitalization. The potential explanation may lie in the fact that despite the weakened virulence of the Omicron strain, the underlying diseases in patients can be aggravated by the Omicron infection, resulting in a higher hospitalization rate, severer or life-threatening illnesses.^2^ Therefore, in preventing the Omicron epidemic, due attention should focus on the population with underlying diseases. Adequate measures, such as vaccination, priority healthcare, and early intervention for primary conditions, should be taken to minimize the hospitalization risk and spare the healthcare system from overwhelming demands.

In the current study, the overall time of nucleic acid negativization (NAN) in the Omicron group was much shorter than that of the Delta group.^14^ This difference may be attributed to the adoption of a booster vaccine in the Omicron group and the recent policy adjustments,^16^ in which the CT value of NAN standard was adjusted from greater than 40 to greater than 35, resulting in an early discharge, in that no infectious viral particles are detected in patients with a CT value of 34 or over^25^ and the measures adopted in containing the local surges in China have demonstrated that the lowered NAN standard has not aggravated the epidemic and has shortened the quarantine time and eased the demands of medical resources.

Ever since the pandemic, vaccination has long been controversial in the elderly population, who are more prone to underlying conditions and thus resultant impaired immunity. This very group of people are particularly reluctant to accept vaccination due to concerns over the side effects. In the current study, further analyses found that the time for NAN was positively correlated with female gender, advanced age and disease severity, but negatively with vaccination, in a dose-dependent manner. This finding is of particular significance for the elderly population, who may have great difficulties in viral clearance due to the underlying diseases and impaired immunity.^26^ They may benefit enormously from active vaccination,^27^ which can stimulate the production of neutralizing antibodies and promote viral clearance. Recent evidence demonstrates the safety and good tolerance of vaccines in the elderly with primary conditions^28^ and a necessity of vaccinating the elderly^29^. In addition, the overall NAN median time of 15 days suggests that it is necessary to quarantine the infected people for at least 15 days in order to minimize the possibility of transmission, which may be prolonged for elderly people with primary conditions. The policy of a five-day home quarantine plus a mask for additional five days, which was once implemented in the United States, has turned out quite inadequate and ineffective in inhibiting the viral transmission.^30^

Altogether, the findings from the current study evidence that vaccination reduces disease severity and accelerates viral clearance, which signifies an urgency of vaccinating the elderly population with underlying illnesses to lower the risks of hospitalization. The median time of NAN suggests that the infected people should be quarantined for at least 15 days to minimize the transmission of the virus.

### Limitations

Firstly, some statistical deviations are inevitable due to the limited patient population and the presence of objective confounding factors such as different vaccination doses and varied sample sizes between the Omicron and Delta groups. Secondly, for an imbalance and deviation may be present in analyzing the hospitalization risks of patients when the asymptomatic and mild patients in the Omicron group were randomly selected to compare with the patients of the moderate category. Finally, the COVID-19 antibody was not detected in the enrolled population of this study, so it is impossible to study the impacts of vaccination on the status of natural immunity and virus clearance.

## Supporting information

appendix with figureS1-2 and tableS1-5

## Data Availability

All data that support the findings of this study are available in the supplementary material of this article.

## Contributors

Hong Li, Baosong Xie, and Xiaoping Chen were responsible for funding acquisition. Hongru Li, Hong Li and Zhijun Su were responsible for design conception. Xiaoping Chen and Yuejiao Ji were responsible for formal analysis. Jiarong Lin, Yanqin Deng, Wen Zhong, Qing Li, Jiabin Fang, Xiaojie Chen, Fang Su, Rong Lin performed data curation. Xiongpeng Zhu, Yi zhang, Zhijun Su, HaiBin Lin and Yu Huang were responsible for resources. Jiarong Lin and Rongguo Yu, Xin Qian verified the underlying data. Hongru Li, Wen Zhong, Qing Li and Jiabin Fang prepared the original draft.

All authors contributed to manuscript reviewing and editing. Jiarong Lin, Xiongpeng Zhu, Yi Zhang, Zhijun Su and Haibin Lin had access to the raw data. All authors had access to all data in the study and the corresponding author had final responsibility for the decision to submit the paper for publication.

## Conflict of Interest

The authors declared no interest conflicts.

## Acknowledgements

This work was supported by the following Fundings:Central Government Guiding Local Science and Technology Development of Fujian Province(2021L3018). Fujian Science and Technology Guidance Project(2021Y0100), The Major Health Research Project of Fujian Province(2021ZD01001),Natural Science Foundation of Fujian Province (2021J01658 、2019J01178), We thank Professor Hongzhi Huang from Fujian Medical University for proofreading and polishing the manuscript.

## Notes

### Competing Interest Statement

The authors have declared no competing interest.

### Author Declarations

Informed consent was granted by all patients and the study protocol was approved by the Ethics Committee of the First Hospital of Quanzhou City (No. 202212) and of the Affiliated Hospital of Putian University (No. 202152).

