## appendix with figureS1-2 and tableS1-5 for "The effects of vaccination on the disease severity and factors for viral clearance and hospitalization in Omicron-infected patients :A retrospective observational cohort study from recent regional outbreaks in China"

**Table of Contents**

**Supplementary Methods**	2

Inclusion Criteria	2

Statistical analysis main script	2

**Figure S1.** The flow chart of the study design	6

**Table S1.** Test kits and apparatus used for the Omicron infection in Quanzhou surge	7

**Table S2.** Normal value of blood test results	8

Table S3. The effects of the 7 categories of different vaccination doses and intervals on the disease severity of the Omicron group	9

Table S4. The effects of the 5 categories of different vaccination doses and intervals on the disease severity of the Omicron group	10

Table S5. Analysis of hospitalization risk factors in Delta infection	11

FigureS2 Comparison of clinical features between the Omicron and Delta group	**12**

**Supplementary Methods**

**Inclusion Criteria**

All cases with positive nucleic acid testing were enrolled from the recent local SARS-CoV-2 surges from Fujian, including 3,265 cases of Omicron variant (BA.2; the Omicron group) in Quanzhou from March 13th, 2022 to May 6th, 2022 and 226 cases of Delta variant (B.1.167.2; the Delta group) in Putian from September 10 to October 20, 2021.

**Statistical analysis main script**

#####Study on Influencing Factors of Nucleic Acid Negative Time#####

###missing value interpolation

w<-read.csv("C:\\Users\\jiyue\\Desktop\\qzxg_data\\zhuanyin_raw.csv",head=TRUE)

str(w)

attach(w)

#install.packages("missForest")

library(missForest)

library(randomForest)

iris.imp <- missForest(w,verbose = TRUE)

#iris.imp$ximp	#data after interpolation

iris.imp$OOBerror	#estimate error

write.csv(iris.imp$ximp,file="C:\\Users\\jiyue\\Desktop\\qzxg_data\\zhuanyin_inter.csv",row.names=F,quote=F)

###linear regression

##read the interpolated dataset

dat<-read.csv("C:\\Users\\jiyue\\Desktop\\qzxg_data\\zhuanyin_inter.csv",header = T,

row.names = NULL)

str(dat)

##factor variable transformation

for (i in names(dat)[c(2,4:9,11,18)]){dat[,i] <- as.factor(dat[,i])}

str(dat)

##unary linear regression

t1<- lm(time~sex,data=dat)

summary(t1)

t2<- lm(time~age,data=dat)

summary(t2)

t3<- lm(time~age_level,data=dat)

summary(t3)

t4<- lm(time~fever,data=dat)

summary(t4)

t5<- lm(time~cough,data=dat)

summary(t5)

t6<- lm(time~Basic.diseases,data=dat)

summary(t6)

t7<- lm(time~vaccination,data=dat)

summary(t7)

t8<- lm(time~class,data=dat)

summary(t8)

t9<- lm(time~IL.6,data=dat)

summary(t9)

t10<- lm(time~CRP,data=dat)

summary(t10)

t11<- lm(time~PCT,data=dat)

summary(t11)

t12<- lm(time~WBC,data=dat)

summary(t12)

t13<- lm(time~LDH,data=dat)

summary(t13)

t14<- lm(time~L,data=dat)

summary(t14)

t15<- lm(time~DDi,data=dat)

summary(t15)

t16<- lm(time~APTT,data=dat)

summary(t16)

t17<- lm(time~Special.treatment,data=dat)

summary(t17)

t18<-lm(time~ORF1lab,data=dat)

summary(t18)

t19<-lm(time~N,data=dat)

summary(t19)

##multiple linear regression

attach(dat)

str(dat)

mod1<-lm(time~sex+age+fever+cough+Basic.diseases+vaccination+class+L+Special.treatment,data=dat)

summary(mod1)

#####Comparison of clinical symptoms between Omicron and Delta#####

#install.packages("forestmodel")

#install.packages("survival")

#install.packages("dplyr")

#install.packages("forplo")

library(forestmodel)

library(survival)

library(dplyr)

library(forplo)

dat<-read.csv("C:\\Users\\jiyue\\Desktop\\qzxg_data\\qvsp.csv",header = T,row.names = NULL)

str(dat)

dat$place<-factor(dat$place,levels=c(0,1), labels=c("Delta","Omicron"))

dat$Fever<-factor(dat$Fever,levels=c(0,1), labels=c("NO","Yes"))

dat$Nasal.congestion<-factor(dat$Nasal.congestion,levels=c(0,1), labels=c("NO","Yes"))

dat$Sore.throat<-factor(dat$Sore.throat,levels=c(0,1), labels=c("NO","Yes"))

dat$Cough<-factor(dat$Cough,levels=c(0,1), labels=c("NO","Yes"))

dat$Expectoration<-factor(dat$Expectoration,levels=c(0,1), labels=c("NO","Yes"))

dat$Fatigue<-factor(dat$Fatigue,levels=c(0,1), labels=c("NO","Yes"))

dat$Dyspnea<-factor(dat$Dyspnea,levels=c(0,1), labels=c("NO","Yes"))

dat$Diarrhea<-factor(dat$Diarrhea,levels=c(0,1), labels=c("NO","Yes"))

dat$Inappetence<-factor(dat$Inappetence,levels=c(0,1), labels=c("NO","Yes"))

dat$Emesis<-factor(dat$Emesis,levels=c(0,1), labels=c("NO","Yes"))

dat$Headache.or.Sore.muscle<-factor(dat$Headache.or.Sore.muscle,levels=c(0,1), labels=c("NO","Yes"))

str(dat)

attach(dat)

###binary logistic regression

mod1<-glm(place~.,data=dat,family="binomial")

summary(mod1)

forest_model(mod1)

###draw forest map

forplo(mod1,

sort=TRUE, #order by OR value

left.align=TRUE,

add.arrow.left=TRUE,

add.arrow.right=TRUE,

shade.every=1,

ci.edge=T,

ci.lwd = 2,

char=20,

col = "darkorange",

right.bar = T,

row.labels=c('Fever',

'Nasal.congestion',

'Sore.throat',

'Cough',

'Expectoration',

'Fatigue','Dyspnea','Diarrhea',

'Inappetence','Emesis',

'Headache.or.Sore.muscle'),

title='Logistic regression, sorted by OR',

save = TRUE,

save.path = "C:\\Users\\jiyue\\Desktop\\qvsp",

save.name = "froplo",

save.type = "png",

save.height = 5.5,

save.width = 7)

#####Percentage plot of stacked columnar#####

#install.packages("ggplot2")

library(ggplot2)

data<-read.csv("C:\\Users\\jiyue\\Desktop\\qzxg_data\\type_class.csv",header=T,

row.names = NULL)

data

##factor sort, set to the order when the table is imported

sorder1 = factor(data$type1,levels=unique(data$type1),order=TRUE)

porder1 = factor(data$class1,levels=unique(data$class1),order=TRUE)

##set font

windowsFonts(A=windowsFont("Times New Roman"),

B=windowsFont("Arial"))

##Taking strain type as abscissa, proportion as ordinate, typing and classification, drawing stacking map

P <- ggplot(data=data,aes(x=sorder1,y=num1,fill=porder1)) +

geom_bar(stat="identity",position="stack") +

labs(x="Variants",y="Proportion",fill="Disease severity")

P+theme(axis.text.x=element_text(angle=0,vjust=1,hjust=0.5,family="A",face="bold",size=15))

**Figure S1.** The flow chart of the study design

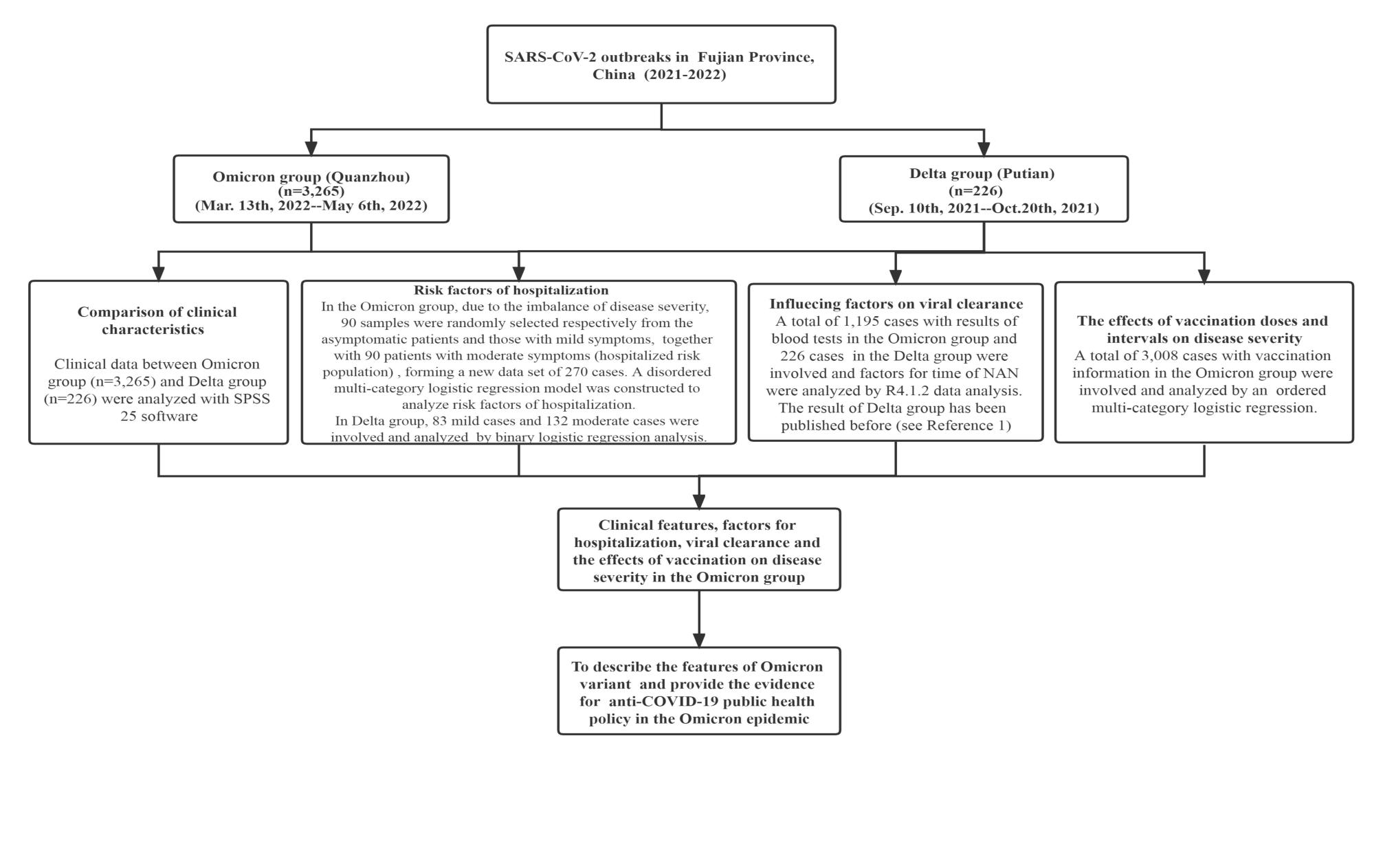

**FigureS1 in appendix：The flow chart of study design**

**NAN:nucleic acid negativization**

**Reference1：**Li H, Lin H, Chen X, et al. Unvaccinated Children Are an Important Link in the Transmission of SARS-CoV-2 Delta Variant (B1.617.2): Comparative Clinical Evidence From a Recent Community Surge. *Frontiers in cellular and infection microbiology* 2022; **12**: 814782.

**Table S1.** Test kits and apparatus used for the Omicron infection in Quanzhou surge

| **Number** | **Test Item** | **Apparatus** | **Kit** |
| --- | --- | --- | --- |
| 1 | Blood routine test | SYSMEX, XN-10 or XN-20, Japan | Corollary Reagent |
| 2 | Biochemistry analysis | BECKMAN, AU5800, American | Corollary Reagent |
| 3 | CRP | PUMEN ,PA990, China | Corollary Reagent |
| 4 | Humoral immunity test | BECKMAN,IMMGE800, American | Corollary Reagent |
| 5 | IL-6 | Roch,cobas8000, Switzerland | Corollary Reagent |
| 6 | Urine test | Sysmex, UF-5000, Japan | Corollary Reagent |
| 7 | Blood coagulation function | WERFEN, ACL TOP700, American | Corollary Reagent |
| 8 | Lymphocyte and subsets count | BECKMANCOULTER, NAVIOS , American | BECKMANCOULTER Multitest IMK Kit |
| 9 | Cytokines | BECKMANCOULTER, NAVIOS , American | BECKMANCOULTER Multitest IMK Kit |
| 10 | SARS-Cov-2 RT-PCR | Nucleic Acid Extraction System：BOJIE,SHANGHAI  PCR Amplifier：ABI,Q5,American | Extraction kit： Bioer  Nucleic acid amplification Kit: Shanghai BJ |
| 11 | Microbe testing | BD,PHOENIX100, American | Corollary Reagent |

**Table S2.** Normal value of blood test results

| **Number** | **Test Item** | **Normal value** |
| --- | --- | --- |
| 1 | WBC count(10^9/l) | 3.5-9.5 |
| 2 | Lymphocyte count(10^9/l) | 1.1-3.2 |
| 3 | CRP（mg/l） | ≤8 |
| 4 | PCT（ng/l） | <0.5 |
| 5 | IL-6(pg/ml) | 0-7 |
| 6 | LDH（U/L） | 120-250 |
| 7 | APTT（s） | 24.9-37.6 |
| 8 | Ddi（mg/l） | 0-0.55 |
| 9 | SARS-Cov-2 RT-PCR | >35 |

**Table S3. The effects of the 7 categories of different vaccination doses and intervals on the disease severity of the Omicron group**

|  |  | **Coefficients** | **OR（95%CI）** | **p-value** |
| --- | --- | --- | --- | --- |
| **Threshold** | Asymptomatic | 0.226 | 1.254 | 0.055 |
|  | Mild | 3.185 | 24.167 | 0 |
| **Location** | No vaccination | Reference | | |
|  | <21 days post dose one | -0.504 | 0.604(0.054-6.753) | 0.682 |
|  | ≥21 days post dose one，but no dose two | 0.069 | 1.071(0.696-1.649) | 0.755 |
|  | <21 days post dose two | -1.728 | 0.178(0.021-1.470) | 0.109 |
|  | ≥21 days post dose two，but no dose three | -0.252 | 0.777(0.601-1.006) | 0.056 |
|  | <21 days post dose three | -0.276 | 0.759(0.517-1.113) | 0.158 |
|  | ≥21 days post dose three | -0.481 | 0.618(0.475-0.803) | <0.001 |

Note: The effects of 7 different vaccination doses and intervals on disease severity (with “asymptomatic”, “mild” and “moderate” indicating the increasing disease severity) in the Omicron group were analyzed by multivariate ordered logistic regression.

**Table S4. The effects of the 5 categories of different vaccination doses and intervals on the disease severity of the Omicron group**

|  |  | **Coefficients** | **OR（95%CI）** | **p-value** |
| --- | --- | --- | --- | --- |
| **Threshold** | Asymptomatic | 0.24 | 1.271（1.008-1.605） | 0.043 |
|  | Mild | 3.197 | 24.459（18.047-33.149） | <0.001 |
| **Location** | No vaccination |  |  |  |
|  | <21 days post dose one | -0.49 | 0.613（0.055-6.848） | 0.691 |
|  | ≥21 days post dose one to <21 days post dose two | -0.025 | 0.975（0.638-1.492） | 0.908 |
|  | ≥21 days post dose two to <21 days post dose three | -0.235 | 0.791（0.612-1.020） | 0.071 |
|  | ≥21 days post dose three | -0.467 | 0.627（0.482-0.815） | 0.001 |

Note: The effects of 5 different vaccination doses and intervals on disease severity (with “asymptomatic”, “mild” and “moderate” indicating the increasing disease severity) in the Omicron group were analyzed by multivariate ordered logistic regression.

**Table S5.** Analysis of hospitalization risk factors in Delta infection

|  | **Total regression** | | | **After removing insignificant variants one by one** | | |
| --- | --- | --- | --- | --- | --- | --- |
|  | **Coefficients** | **OR(95%CI）** | **p-value** |  | **OR(95%CI）** | **p-value** |
| （Intercept) | -0.605 |  | 0.150 | -0.388 | —— | 0.159 |
| Age | 0.046 | 1.04（1.01-1.07） | 0.005 ** | 0.041 | 1.04（1.02-1.07） | 0.003 ** |
| Sex | 0.119 | 1.13（0.6-2.09） | 0.714 |  |  |  |
| Fever | 0.082 | 1.17（0.62-2.21） | 0.808 |  |  |  |
| Nasal congestion | -19.021 | —— | 0.996 |  |  |  |
| Sore throat | -19.012 | —— | 0.996 |  |  |  |
| Cough | 0.298 | 1.84（0.83-4.22） | 0.497 | 0.914 | 2.49（1.32-4.87） | 0.006 ** |
| Expectoration | 0.764 | 1.70（0.59-5.09） | 0.190 |  |  |  |
| Fatigue | -0.200 | 0.89（0.44-1.80） | 0.596 |  |  |  |
| Dyspnea | 15.897 | —— | 0.995 |  |  |  |
| Diarrhea | 1.369 | 2.54（0.30-54.18） | 0.287 |  |  |  |
| Inappetence | 15.727 | —— | 0.990 |  |  |  |
| Emesis | -0.554 | 1.63（0.13-38.37） | 0.729 |  |  |  |
| Headache or Sore muscle | 17.007 | —— | 0.987 |  |  |  |
| Underlying conditions | 0.450 | 1.54（0.41-7.55） | 0.611 |  |  |  |
| Vaccination |  |  |  |  |  |  |
| One-dose | 0.710 | 2.01（0.25-42.27） | 0.552 | 0.588 | 1.80（0.23-37.58） | 0.618 |
| Two-dose | -1.122 | 0.36（0.12-1.01） | 0.049 * | -1.066 | 0.34（0.12，0.92） | 0.038 * |

Note: In the Delta group, except few asymptomatic and severe cases, mild level was set as 0 and moderate level (hospitalization risk population） as 1 and a disordered multi-category logistic regression model was constructed for analysis.

**Figure S2. The comparison of Clinical features between the Omicron group and Delta group**

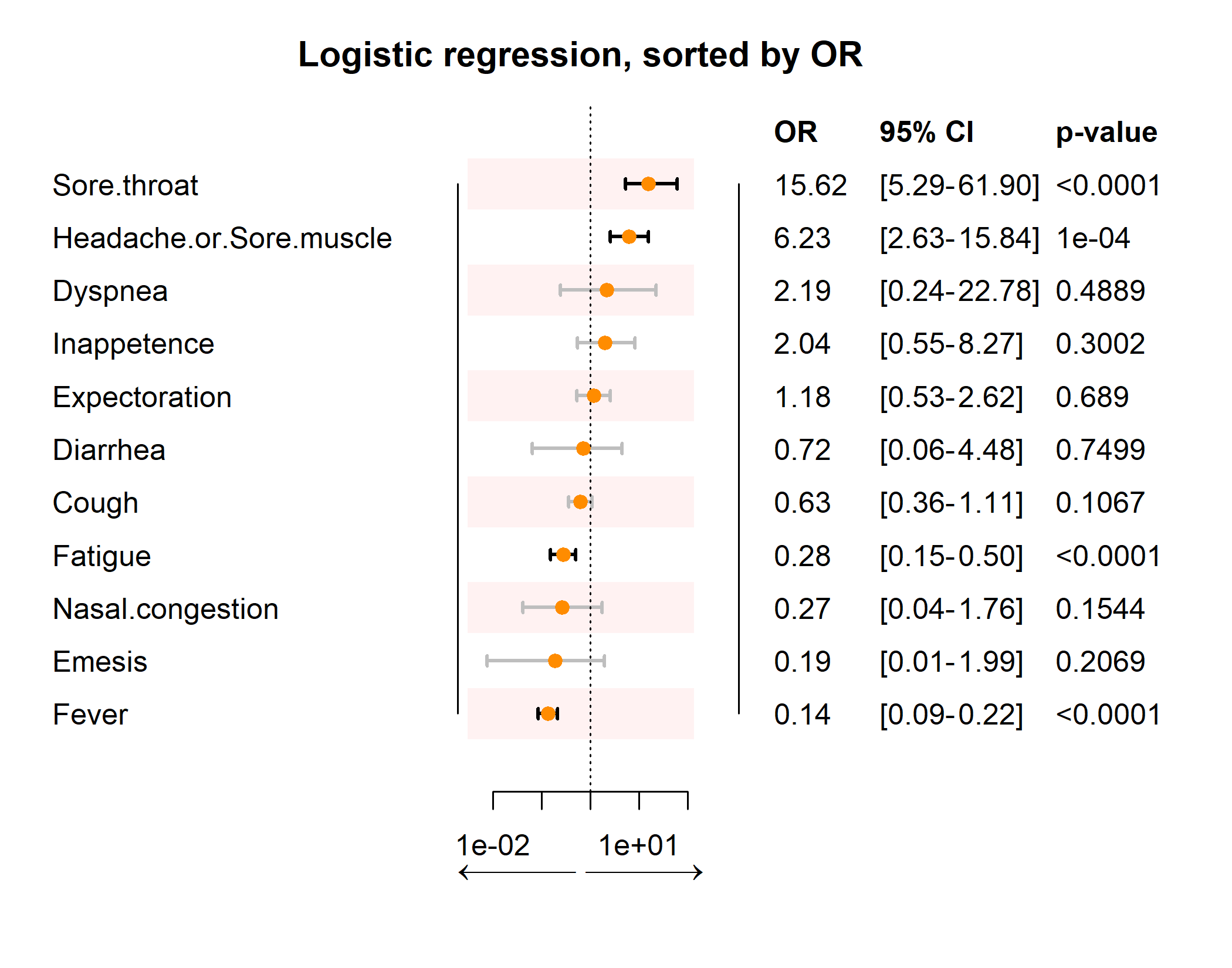

Note: In the Omicron group, a 1:1 sample size (matched with 90 patients with moderate symptoms) were randomly selected from asymptomatic and mild patients, respetively, forming a new data set of 270 cases in total. In the Delta group, with 6 severe and critical cases excluded, 220 cases with asymptomatic, mild, moderate symptoms formed a new data set. Then these two data sets constructed a forest plot by binary logistic regression model. The results indicated that compared with the Delta-infected individuals, the Omicron-infected patients were more likely to present sore throat, headache or myalgia, less likely to experience fatigue and fever (*p*<0.05).
